# Determinants of sleep quality in adults during the COVID-19 pandemic: *COVID-Inconfidentes*, a population-based study

**DOI:** 10.1101/2021.09.29.21264305

**Authors:** Luiz Antônio Alves de Menezes Júnior, Luciano Garcia Lourenção, Amanda Cristina de Souza Andrade, Júlia Cristina Cardoso Carraro, George Luiz Lins Machado-Coelho, Adriana Lúcia Meireles

## Abstract

**Background:** The coronavirus disease 2019 (COVID-19) pandemic has had a negative effect on the health and behavior of the world’s population.

**Objectives:** To evaluate sleep quality and its associated factors in adults during the COVID-19 pandemic in Brazil.

**Methods:** This is a population-based serological survey of 1762 adults collected from October to December 2020 in the Iron Quadrangle region, Brazil. To measure sleep quality, we used the Pittsburgh Sleep Quality Index questionnaire and socio-demographic, health, health related behaviors, anxiety, vitamin D, weight gain/loss, and pandemic characteristics were assessed using a structured questionnaire. Univariate and multivariate analyses were performed to identify the factors associated with sleep quality.

**Results:** More than half of the individuals evaluated had poor sleep quality (52.5%). In multivariate analysis, factors related to sleep quality included living alone (OR=2.36; 95%CI: 1.11-5.00), anxiety disorder (OR=2.22; 95%CI: 1.20-4.14), 5.0% weight loss during the pandemic (OR=1.66; 95%CI: 1.01-2.76), weight gain of 5.0% (OR=1.90; 95%CI: 1.08-3.34), insufficient vitamin D scenario (OR=1.47; 95%CI: 1.01-2.12), and symptoms of COVID-19 (OR=1.94; 95%CI: 1.25-3.01).

**Conclusions:** Our study revealed that more than half of the participants had poor sleep quality during the COVID-19 outbreak, and the factors associated with poor sleep quality were related to the pandemic, such as insufficient vitamin D scenario and weight change.

## Introduction

In late 2019, a novel coronavirus (SARS-CoV-2) was identified as causing pneumonia in Wuhan, China. The infection has spread rapidly worldwide, resulting in a global pandemic with a great impact on the healthcare system of many countries (Guan et al. 2020). Since the beginning of the pandemic to almost two years later, Brazil is among the most affected countries and remains in the top five countries with the highest number of infected people and deaths from COVID-19 (Hannah et al. 2021).

Due to the high propagation of COVID-19 and the fact that little is known about its natural history, control measures have been adopted, such as respiratory hygiene, use of masks, and social restrictions (Brooks et al. 2020). These measures, along with the pandemic scenario, led to drastic changes in the population’s lifestyle, such as reduced physical activity, changes in food intake, sun exposure (Brooks et al. 2020; Di Renzo et al. 2020), and other factors directly related to sleep quality (Barros et al. 2020; Gao et al. 2018; Lancel, Boersma, and Kamphuis 2021).

Sleep is essential for maintaining physiological parameters and plays an important role in hormone release, regulation of cardiovascular activity, and glucose (Irwin 2015). In addition, poor sleep quality, especially if chronic, may adversely affect the immune system components, leading to increased vulnerability to infectious diseases such as COVID-19 (Irwin 2015).

Therefore, considering that a pandemic alters the daily routine and life habits of the population (Flanagan et al. 2021), the present study aimed to evaluate sleep quality and its associated factors during the COVID-19 pandemic.

## Methods

### Study design

A cross-sectional population-based serological study was performed between October and December 2020 in two medium-sized cities located in the central-southern region of the state of Minas Gerais, known as the Iron Quadrangle. It is a metallurgical area that is considered the largest national producer of iron ore. The study design followed the recommendations of the seroepidemiological investigation protocol for SARS-CoV-2 infection by the World Health Organization (WHO 2020a). The research ethics committee of Federal University of Minas Gerais approved the project (certificate of ethics submission Nº. 32815620.0.1001.5149). All procedures adopted in this study followed the Declaration of Helsinki and the Brazilian guidelines and standards for research involving humans, and written informed consent was collected from participants.

The survey was carried out in three stages, with intervals of 21 days, considering the incubation period of the virus SARS-CoV-2, in which different census sectors were evaluated in each city (WHO 2020a). The sample size calculation was based on the population estimate for each city, taking into consideration a confidence level of 95%, a design effect equal to 1.5, and the parameters presented in a previous study (Meireles et al. 2021). A three-stage conglomerate sampling design was adopted: census sector (selected for each stage, randomly and without replacement), households (selected by a systematic sampling process), and residents (one resident was selected randomly, using the Sorteador de Nomes® applicative). The sample weight of each selected unit (census tract, household, and individual) was calculated and adjusted to compensate for the loss of interviews due to non-response, and the weight of the household and the selected resident was calibrated(Meireles et al. 2021).

### Data collection

The data collection process included listing and approaching households, recruiting participants, collecting venous blood, and conducting interviews. The activities were carried out during weekends (Friday, Saturday, and Sunday), aiming to enhance the participation of residents who worked during the week and thus increasing the representativeness of this population group.

During face-to-face interviews conducted by trained interviewers, a questionnaire containing sociodemographic, health conditions, pandemics, and sleep information was administered. The socio-demographic variables evaluated were sex, age (18-34, 35-59, or ≥ 60 years), marital status (married or unmarried), living status (living alone or not alone), education (nonliterate, up to 9 years, or > 9 years), family income (≤ 2, 2-4, or > 4 times the minimum wage), worker (yes or no), and current shift work (yes or no). Furthermore, we also evaluated work from home schedule, in “no work from home,” “partial work from home,” (some days at home and some at the workplace), and “total work from home,” (all work activities performed from home). Responses were dichotomized as “no work from home” and “yes, (partial or fulltime work from home”).

Health conditions included self-reported chronic diseases, measured by the following question: “Has a physician or other health professional said that you have: high blood pressure, diabetes, asthma, lung disease, chronic kidney disease, depression, anxiety disorder, cancer, heart disease, or thyroid disease?”. Individuals were assessed for a separate chronic disease (yes or no) and also combined into two categories: with morbidity (reporting at least one disease) and without morbidity (no disease). Individuals were also assessed for chronic physical pain (physical pain present for 3 months or more), current smoking (yes or no), current alcohol drinking (yes or no), physical activity (inactivity, or activity at least 150– 300 minutes of moderate-intensity aerobic physical activity, or at least 75–150 minutes of vigorous-intensity aerobic physical activity per week) (WHO 2020b), and self-rated health were also assessed. The last variable was measured by the question “In general, would you say that your health is: very good, good, fair, poor, or very poor?”. The answers were dichotomized as “poor (fair, poor, very poor)” and “good (very good, good).”

We also assessed nutritional status according to body mass index (BMI). Weight and height were self-reported and used to calculate the BMI and classified as underweight (BMI < 18.5 kg/m^2^ if < 60 years or BMI < 22.0 kg/m^2^ if ≥ 60 years), eutrophic (BMI 18.5-24.9 kg/m^2^ if < 60 years or BMI 22.0-27.0 kg/m^2^ if ≥ 60 years), and overweight (BMI ≥ 25.0 kg/m^2^ if < 60 years or BMI ≥ 27.0 kg/m^2^ if ≥ 60 years) (Organização Pan-Americana de Saúde 2001; WHO 1995).

We also assessed the change in their weight during the pandemic, for which the participants were asked about their weight before the pandemic and their current weight. The change was classified into three groups according to the percentage of weight change, with “no weight change” if -5.0% to 5.0%, “weight loss” if -5.0% or more, and “weight gain” if 5.0% or more during the pandemic.

The average daily sun exposure was evaluated and classified as “insufficient” when less than 30 min/day and “sufficient” when greater than or equal to 30 min/day (Holick 2007). We also evaluated a possible scenario of vitamin D adequacy, considering the extent of the time of sun exposure and the consumption of food supplements that are sources of vitamin D. Thus, we classified the vitamin D scenario as sufficient when the average daily sun exposure was greater than or equal to 30 min/day or the individual reported consuming a food supplement that is a source of vitamin D (Holick 2007).

Questions related to the COVID-19 pandemic were also evaluated, such as presenting at least one symptom in the last 15 days (fever, sore throat, cough, dyspnea, tachycardia, diarrhea, vomiting, anosmia, ageusia, and fatigue); social restriction since the beginning of the pandemic; a family member who is in the COVID-19 risk group (people over 60 years or with cardiovascular disease, diabetes, respiratory disease, neurological or renal disease, immunosuppression, obesity, asthma, pregnant women, or women who have had children less than 42 days before); and pandemic period (7-8.5 months, and 8.5-9 months after the beginning of the pandemic in Brazil, March 2020). Further, we asked about their daily routine activities during the pandemic, measured with the question “How are your routine activities during the pandemic of COVID-19?”: “Left home frequently to work, study, or other regular activity” (yes or no); “Go outside, on the streets, for exercise” (yes or no); and “Go to the gym to do some physical activity” (yes or no).

### Measurement of sleep quality

The Pittsburgh Sleep Quality Index (PSQI) questionnaire was used to measure the sleep quality of the study participants. This instrument is composed of 19 questions categorized into seven components: subjective sleep quality (C1), sleep latency (C2), sleep duration (C3), habitual sleep efficiency (C4), sleep disturbances (C5), use of sleep medication (C6), and daytime dysfunction (C7). The sum of the scores produces an overall score ranging from 0 to 21, where the highest score indicates the worst sleep quality. An overall score greater than five indicates major difficulties in at least two components, or moderate difficulties in more than three components (Buysse et al. 1989). The Brazilian version of the Pittsburgh Sleep Quality Index had an overall reliability coefficient (Cronbach’s α) of 0.82, indicating a high degree of internal consistency (Bertolazi et al. 2011).

In this study, sleep quality was classified as good quality (PSQI score ≤ 5) and poor quality (PSQI > 5). Abnormal sleep in a specific domain was defined as a score of ≥ 2.

### Statistical analysis

Statistical analyses were performed considering the complex design of the sample using the svy command of Stata® software, version 15.0. Data are presented as percentages and confidence intervals (95% CI). Data were compared using the chi-square test. Univariate and multivariate logistic regression analyses were used to determine the association between sleep quality and sociodemographic factors, health conditions, and COVID-19 related variables. Independent variables that had an association at a p-value of 0.2 were used in multivariate logistic regression with a stepwise backward elimination procedure controlling for the pandemic period variable.

In addition, to verify a possible effect modification on sleep quality, a bivariate analysis was performed on the multivariate model of the interaction between the associated factors.

## Results

### Characteristics of study participants

Table 1 shows the sociodemographic characteristics of the study participants according to sleep quality (overall PSQI score > 5 vs. ≤ 5). Of the participants, 51.9% were women, the most prevalent age group was 35-59 years (45.6%), most were married (53.2%), had more than 9 years of schooling (68.8%), and had a family income of less than or equal to two times minimum wages (41.1%). Regarding work-related variables, 52.5% of the evaluated individuals were active workers, 38.6% were working from home, and 8.6% worked alternate shifts.

**Table 1.**
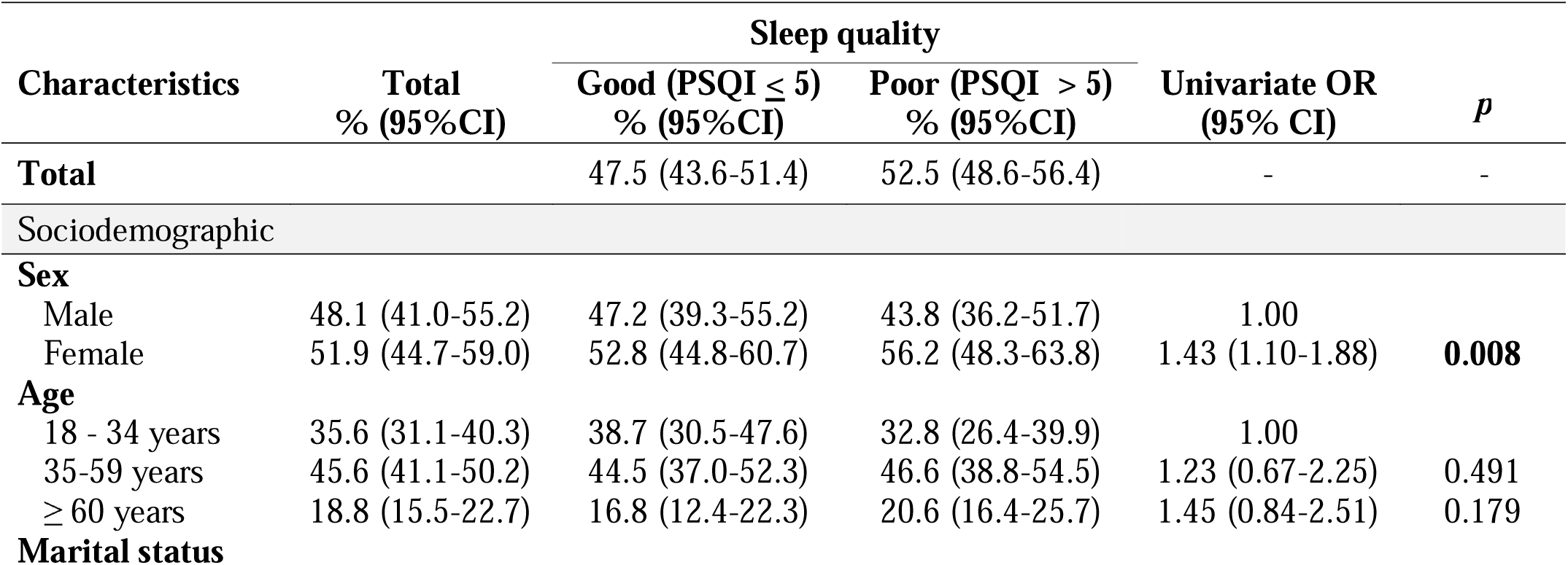
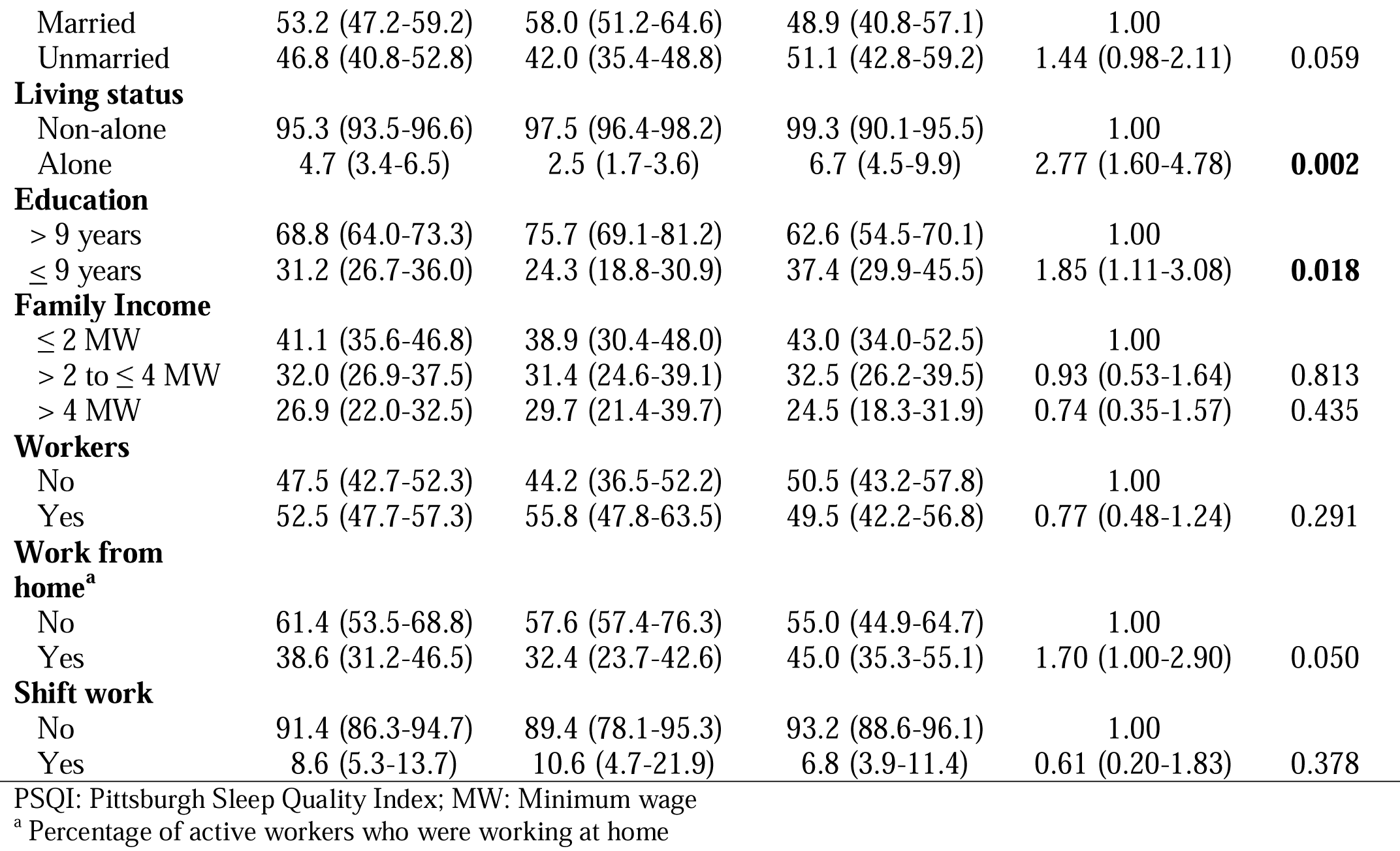
Sociodemographic characteristics according to sleep quality during pandemic

Concerning their health conditions, more than half of the participants had at least one chronic disease (52.3%), consumed alcohol (58.2%), were physically inactive (69.2%), and overweight (61.4%) (Table 2). At least 12% of individuals experienced a 5.0% weight loss or gain during the pandemic (12.4% and 17.7%, respectively), 35.0% had a daily sun exposure of less than 30 min, and 27.1% had an insufficient vitamin D scenario. Regarding the COVID-19 variables, 28.6% reported at least one symptom of COVID-19 in the 15 days before the survey, 85.4% reported that they had experienced social restrictions since the start of the pandemic, and 59.2% had at least one family member in the risk group for severe COVID-19 (Table 3).

**Table 2.**
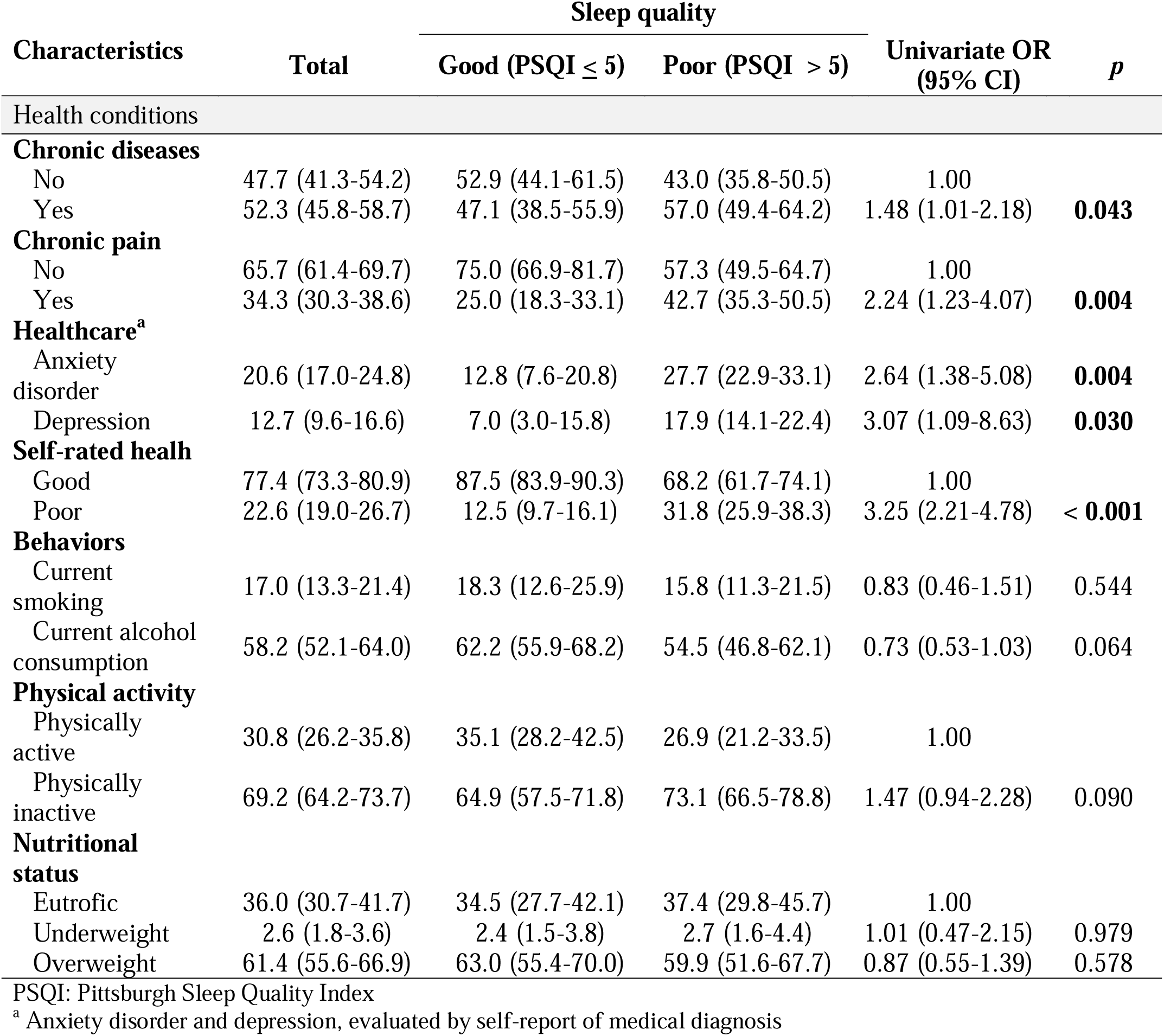
Health conditions according to sleep quality during pandemic

**Table 3.** COVID-19 related variables according to sleep quality during pandemic

| Characteristics | Total | Sleep quality |  | Univariate OR<br>(95% CI) | *p* |
| --- | --- | --- | --- | --- | --- |
|  |  | Good<br>(PSQI ≤ 5) | Poor<br>(PSQI > 5) |  |  |
| **Weight change<sup>a</sup>** |  |  |  |  |  |
| Δ -5% to +5% | 69.9 (64.8-74.5) | 76.6 (71.6-80.9) | 63.9 (56.5-70.8) | 1.00 |  |
| Δ ≤ -5% | 12.4 (9.3-16.4) | 10.1 (7.3-13.7) | 14.5 (9.7-21.0) | 1.72 (1.02-2.91) | **0.043** |
| Δ ≥ +5% | 17.7 (14.8-21.1) | 13.3 (9.9-17.8) | 21.6 (16.7-27.5) | 1.93 (1.17-3.18) | **0.010** |
| **Exposure sun** |  |  |  |  |  |
| ≥ 30 min/day | 64.5 (59.3-70.3) | 69.1 (62.6-74.8) | 61.3 (53.7-68.4) | 1.00 |  |
| < 30 min/day | 35.0 (29.7-40.7) | 30.9 (25.1-37.4) | 38.7 (31.6-46.3) | 1.41 (1.01-1.98) | **0.049** |
| **Vitamin D supplementation** |  |  |  |  |  |
| No | 77.9 (73.3-81.9) | 80.2 (74.6-84.8) | 75.9 (69.4-81.4) | 1.00 |  |
| Yes | 22.1 (18.0-26.7) | 19.8 (15.2-25.4) | 24.1 (18.6-30.6) | 1.28 (0.84-1.95) | 0.237 |
| **Vitamin D scenario<sup>b</sup>** |  |  |  |  |  |
| Sufficient | 72.9 (68.1-77.3) | 76.7 (72.3-81.1) | 69.3 (62.3-75.4) | 1.00 |  |
| Insufficient | 27.1 (22.7-31.9) | 23.0 (18.9-27.7) | 30.7 (24.5-37.7) | 1.48 (1.07-2.04) | **0.017** |
| **SARS-CoV-2** |  |  |  |  |  |
| Soronegative | 94.8 (93.0-96.2) | 94.9 (91.8-96.9) | 94.7 (92.1-96.5) | 1.00 |  |
| Soropositive | 5.2 (3.8-7.0) | 5.1 (3.1-8.2) | 5.3 (3.5-7.9) | 1.03 (0.51-2.06) | 0.927 |
| **Symptoms of COVID-19** |  |  |  |  |  |
| No | 71.4 (66.7-75.8) | 79.8 (75.0-83.8) | 63.8 (56.7-70.4) | 1.00 |  |
| Yes | 28.6 (24.2-33.3) | 20.2 (16.2-25.0) | 36.2 (29.6-43.3) | 2.23 (1.56-3.20) | **< 0.001** |
| **Risk group in family** |  |  |  |  |  |
| No | 40.8 (33.8-48.2) | 46.2 (38.8-53.7) | 36.0 (28.2-44.5) | 1.00 |  |
| Yes | 59.2 (51.8-66.2) | 53.8 (46.3-61.2) | 64.0 (55.5-71.8) | 1.53 (1.17-1.99) | **0.002** |
| **Pandemic period** |  |  |  |  |  |
| 8.5-9 months | 18.9 (14.6-24.1) | 22.5 (16.6-29.6) | 15.6 (11.7-20.6) | 1.00 |  |
| 7-8.4 months | 81.1 (75.9-85.4) | 77.5 (70.4-83.3) | 84.4 (79.4-88.3) | 1.56 (1.09-2.23) | **0.015** |
| **Daily routine in pandemic** |  |  |  |  |  |
| Social contact restriction | 62.6 (58.3-66.7) | 56.7 (48.3-64.7) | 68.0 (61.3-73.9) | 1.62 (0.97-2.71) | 0.066 |
| Physical activity in the street | 23.9 (20.3-28.0) | 26.1 (18.8-34.9) | 22.0 (18.1-26.5) | 0.80 (0.46-1.38) | 0.419 |
| Physical activity in the gym | 10.2 (6.7-15.2) | 14.2 (8.5-22.6) | 6.6 (3.7-11.4) | 0.42 (0.19-0.91) | **0.028** |

PSQI: Pittsburgh Sleep Quality Index
<sup>a</sup> Weight change during the pandemic, from self-reported weight<sup>b</sup> Sufficient: Sun exposure > 30 min/day or vitamin D supplements; Insufficient: Sun exposure < 30 min/day and no vitamin D supplements

The distribution of the PSQI scores and subdomains is presented in Figure 1.

**Fig. 1:**
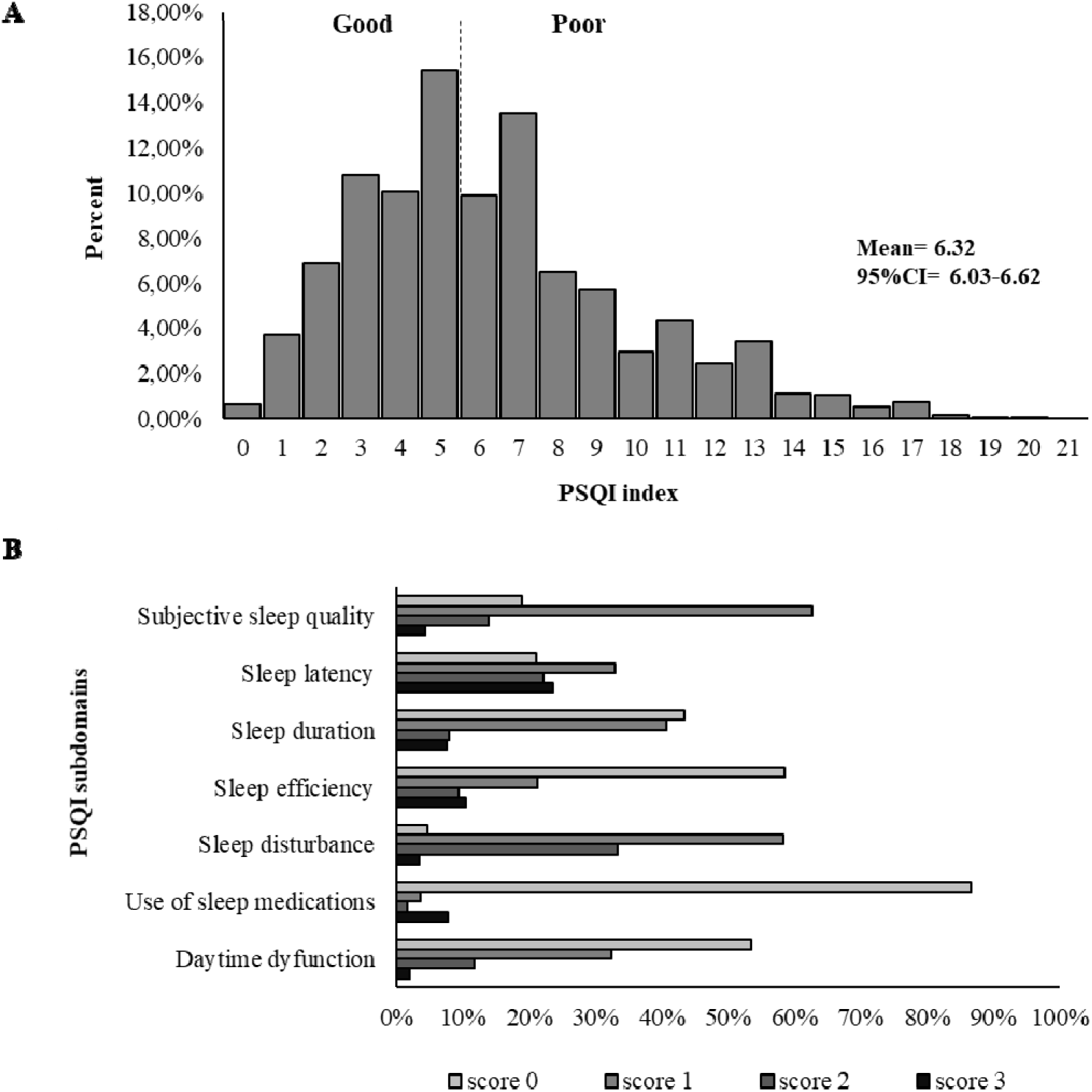
Frequency distribution of PSQI score (A) and domain scores (B) of sleep quality during the COVID-19 pandemic

The PSQI had a mean score of 6.32 (95% CI 6.03-6.62), and the prevalence of poor sleep quality was reported by 52.5% of the total sample (43.8% men, and 56.2% women) (p=0.008).

### Distribution of poor sleep quality and subdomains

Table 4 shows the distribution of the abnormal Pittsburgh sleep quality subdomains by age group and sex. The prevalence rates of abnormal specific sleep domains were 18.3% for subjective sleep quality, 45.8% for sleep latency, 15.7% for sleep duration, 20.1% for sleep efficiency, 36.8% for sleep disturbance, 9.6% for use of sleep medications, and 13.9% for daytime dysfunction. Women reported a higher prevalence of abnormal Pittsburgh sleep subdomains for subjective sleep quality, sleep efficiency, and use of sleep medications (p < 0.05).

**Table 4.** Distributions of abnormal Pittsburgh Sleep Quality Index subdomains by sleep quality, age and sex.

|  | Abnormal Pittsburgh Sleep Quality Index subdomains<sup>a</sup>, n(%) |  |  |  |  |  |  |
| --- | --- | --- | --- | --- | --- | --- | --- |
|  | Subjective sleep quality | Sleep latency | Sleep duration | Sleep efficiency | Sleep disturbance | Use of sleep medications | Daytime dysfunction |
| **Total sample** | 18.3 (14.9-22.4) | 45.8 (41.6-50.1) | 15.7 (12.3-19.8) | 20.1 (16.7-24.1) | 36.8 (32.0-41.9) | 9.6 (7.6-12.1) | 13.9 (11.0-17.5) |
| **Sex** |  |  |  |  |  |  |  |
| Male | 36.6 (27.2-47.2) | 43.8 (34.6-53.5) | 44.3 (35.2-53.7) | 37.5 (28.7-47.1) | 42.8 (32.9-53.2) | 29.0 (19.9-40.3) | 39.8 (27.7-53.3) |
| Female | 63.4 (52.8-72.8) | 56.2 (46.5-65.4) | 55.7 (46.3-64.8) | 62.5 (52.9-71.3) | 57.2 (46.8-67.1) | 71.0 (59.7-80.1) | 60.2 (46.7-72.3) |
| *p-value* | **0.043** | 0.108 | 0.448 | **0.032** | 0.070 | **0.001** | 0.222 |
| **Age** |  |  |  |  |  |  |  |
| 18 - 34 years | 35.6 (25.6-47.7) | 36.0 (29.0-43.6) | 33.7 (22.0-47.8) | 31.7 (22.4-42.8) | 28.8 (21.0-38.1) | 11.1 (5.6-20.9) | 50.2 (38.0-62.3) |
| 35-59 years | 47.0 (35.3-59.0) | 45.3 (37.2-53.7) | 44.8 (32.4-57.8) | 44.8 (36.8-53.0) | 48.2 (37.7-58.9) | 54.4 (43.2-65.1) | 35.0 (25.2-46.3) |
| ≥ 60 years | 17.1 (12.4-23.1) | 18.7 (14.9-23.0) | 21.5 (15.2-29.6) | 23.5 (18.1-30.0) | 23.0 (17.0-30.2) | 34.5 (24.8-45.5) | 14.8 (9.7-21.9) |
| *p-value* | 0.854 | 0.971 | 0.746 | 0.385 | 0.157 | **< 0.001** | **0.004** |

<sup>a</sup>Score for each domain ranges from 0 to 3 (no difficulty to severe difficulty), and a domain score $\geq 2$ indicates abnormal sleep in the domain

In the age subgroups, there was a significant difference in the prevalence of Pittsburgh abnormal sleep subdomains for sleep medication use and daytime dysfunction. Sleep medication use increased with increasing age, and daytime dysfunction was higher in the younger age groups (p < 0.05) (Table 4).

### Associated factors of poor sleep quality

Table 5 shows the association between poor sleep quality and sociodemographic, health conditions, and pandemic variables. In the multivariate model, the following factors were significantly associated with poor sleep quality: living alone (OR=2.36; 95%CI: 1.11-5.00), anxiety disorder (OR=2.22; 95%CI: 1.20-4.14), 5.0% weight loss (OR=1.66; 95%CI: 1.01-2.76), and gain of 5.0% weight during the pandemic (OR=1.90; 95%CI: 1.08-3.34), insufficient vitamin D scenario (OR=1.47; 95%CI: 1.01-2.12), and symptoms of COVID-19 (OR=1.94; 95%CI: 1.25-3.01).

**Table 5.**
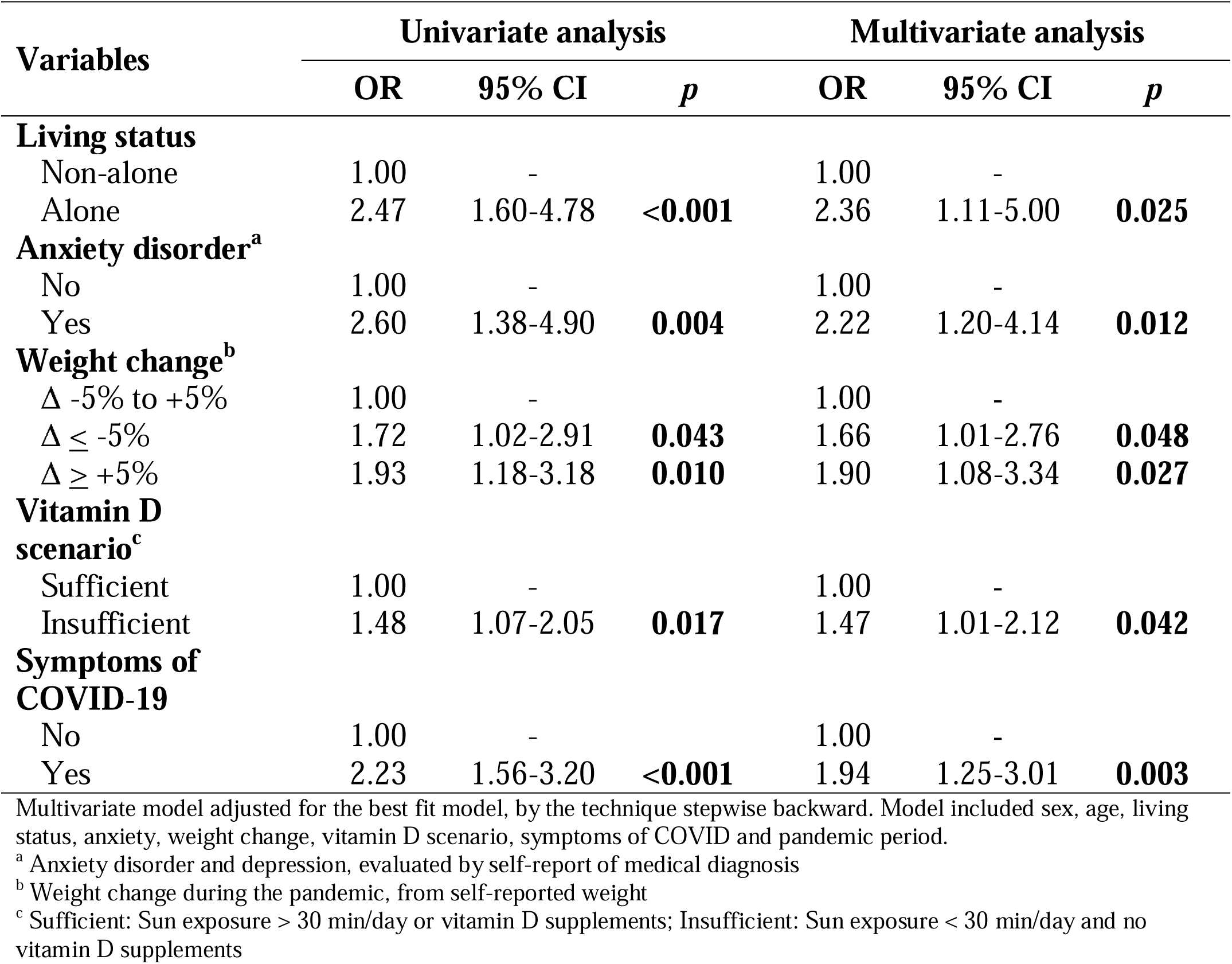
Multivariate logistic regression analysis of factors associated with poor sleep quality.

| Variables | Univariate analysis |  |  | Multivariate analysis |  |  |
| --- | --- | --- | --- | --- | --- | --- |
|  | OR | 95% CI | *p* | OR | 95% CI | *p* |
| **Living status** |  |  |  |  |  |  |
| Non-alone | 1.00 | - |  | 1.00 | - |  |
| Alone | 2.47 | 1.60-4.78 | **<0.001** | 2.36 | 1.11-5.00 | **0.025** |
| **Anxiety disorder<sup>a</sup>** |  |  |  |  |  |  |
| No | 1.00 | - |  | 1.00 | - |  |
| Yes | 2.60 | 1.38-4.90 | **0.004** | 2.22 | 1.20-4.14 | **0.012** |
| **Weight change<sup>b</sup>** |  |  |  |  |  |  |
| $\Delta$ -5% to +5% | 1.00 | - | | 1.00 | - | |
| $\Delta \leq -5\%$ | 1.72 | 1.02-2.91 | **0.043** | 1.66 | 1.01-2.76 | **0.048** |
| $\Delta \geq +5\%$ | 1.93 | 1.18-3.18 | **0.010** | 1.90 | 1.08-3.34 | **0.027** |
| **Vitamin D scenario<sup>c</sup>** |  |  |  |  |  |  |
| Sufficient | 1.00 | - |  | 1.00 | - |  |
| Insufficient | 1.48 | 1.07-2.05 | **0.017** | 1.47 | 1.01-2.12 | **0.042** |
| **Symptoms of COVID-19** |  |  |  |  |  |  |
| No | 1.00 | - |  | 1.00 | - |  |
| Yes | 2.23 | 1.56-3.20 | **<0.001** | 1.94 | 1.25-3.01 | **0.003** |

Multivariate model adjusted for the best fit model, by the technique stepwise backward. Model included sex, age, living status, anxiety, weight change, vitamin D scenario, symptoms of COVID and pandemic period.
<sup>a</sup> Anxiety disorder and depression, evaluated by self-report of medical diagnosis
<sup>b</sup> Weight change during the pandemic, from self-reported weight
<sup>c</sup> Sufficient: Sun exposure > 30 min/day or vitamin D supplements; Insufficient: Sun exposure < 30 min/day and no vitamin D supplements

Based on the factors associated with sleep quality, which were obtained in the adjusted model presented above (Table 5), a chance modification analysis for poor sleep quality was performed assuming the presence of combined changes in these variables (Figure 2). In general, we observed that the variables assessed had a gradient of probability for sleep quality, with the odds of having poor sleep quality increasing when two concurrently altered variables were analyzed. The worst scenario was the concurrence between symptoms of COVID-19 and weight loss (OR= 6.40; 95%CI: 2.00-6.40). Only the variable weight loss when evaluated concomitantly with the insufficient vitamin D scenario variable was not significant (OR, 1.17; 95%CI: 0.47-2.91).

**Fig. 2:**
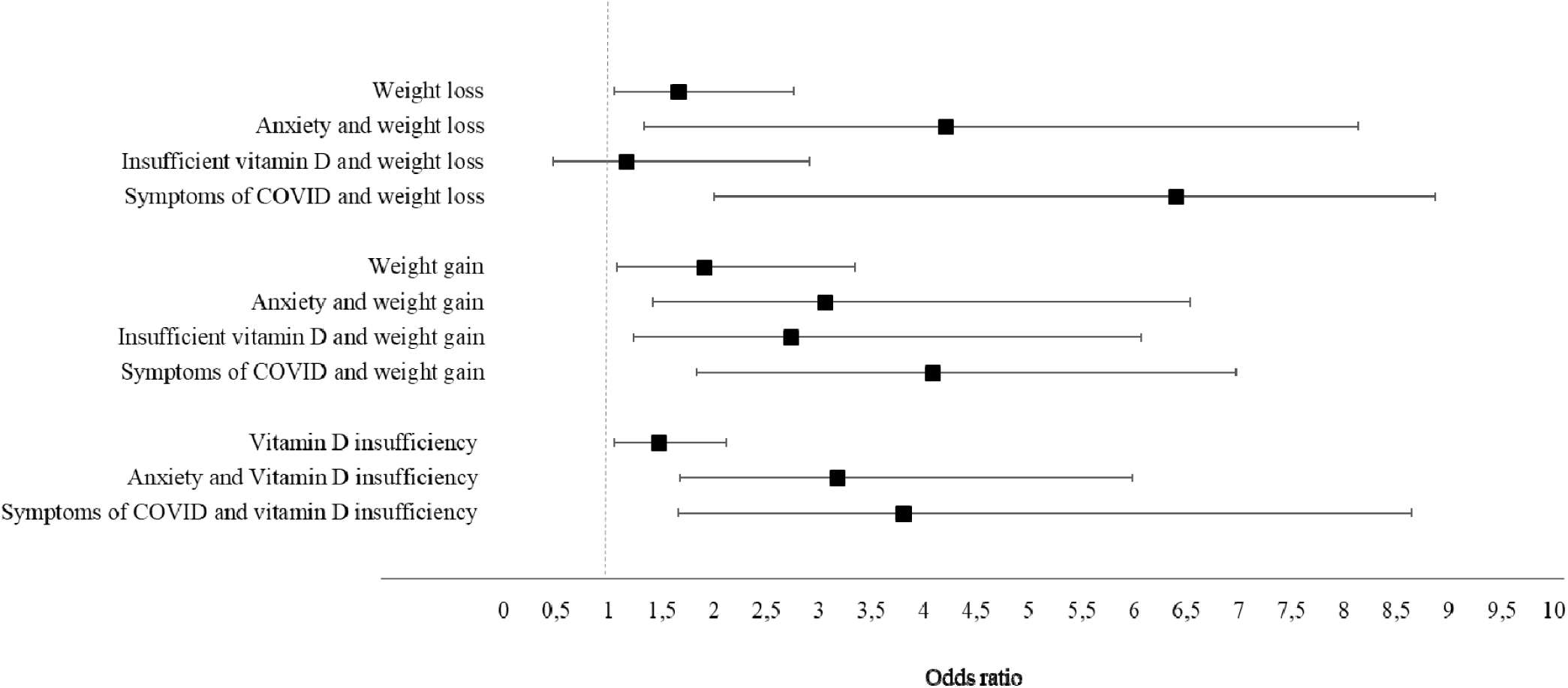
Bivariate association adjusted for weight change, and vitamin D insufficiency with individual parameters associated with poor sleep quality during the COVID-19 pandemic

## Discussion

The current study investigated the prevalence of poor sleep quality and associated factors during the COVID-19 pandemic. We found that more than half of the population had poor sleep quality. Living alone, anxiety disorder, weight change during the pandemic, insufficient vitamin D scenario, and symptoms of COVID-19 increased the chances of having sleep problems.

The most affected PSQI subdomains were sleep latency, sleep disturbance, and sleep efficiency. These results are corroborated by other population studies, in which the perception of poor sleep quality is common, such as a study by the National Sleep Foundation in 2014, which showed that at least 35% of adult Americans considered their sleep to be of poor quality (Knutson et al. 2017). In Brazil, population studies have demonstrated high percentages of individuals with poor sleep quality, especially among women aged 40 to 50 years, without occupation, physically inactive, and with a greater number of health problems (Szwarcwald et al. 2021). A national survey conducted in April 2012 with individuals from 132 different cities found that 76% of the population had at least one sleep complaint, indicating that approximately 108 million Brazilians may be affected by sleep disorders (Hirotsu et al. 2014).

During the pandemic, several factors may contribute to altering the normal architecture of sleep, and population studies are important because they allow us to evaluate how the health outcomes affect the lives of the population. However, few studies with this methodology using the PSQI were conducted during the pandemic, which makes it difficult to compare the results. Our study, conducted from October to December 2020, found a higher prevalence than studies conducted at the beginning of the pandemic, as shown by Krishnamoorthy et al. (2020) in a systematic review, in which approximately 36% of the general population had poor sleep quality, and among healthcare workers, one of the most affected groups during the pandemic, 43% had poor sleep quality (Krishnamoorthy et al. 2020). In Brazil, a study of 45,161 individuals from April to May 2020 showed that during the pandemic, 43.5% (95%CI 41.8;45.3) reported the onset of sleep problems and 48.0% (95%CI 45.6;50.5) had a previous sleep problem that worsened after the pandemic (Szwarcwald et al. 2021). However, it should be noted that this study was conducted online, which usually evaluates a more educated and higher-income group of the population and is different compared to the household survey.

During the pandemic, online tasks have made the workday never-ending and affect the quality of sleep. This work schedule can also reduce the sun exposure of individuals, an important factor since it is the main source of endogenous production of vitamin D (Holick 2007). Here, we assessed whether the vitamin D scenario was associated with sleep quality during a pandemic. We found that individuals with an insufficient vitamin D level were 47% more likely to have poor sleep quality. This association may be explained by the intracellular distribution of vitamin D receptors in areas of the brain that regulate the sleep-wake cycle or through pro-inflammatory mediators. Vitamin D is also involved in the production of melatonin, an essential hormone in the regulation of circadian rhythms and sleep. Melatonin synthesis is controlled by the active form of vitamin D (1,25(OH)D) from the enzyme tryptophan hydroxylase (Romano et al. 2020). This suggests a possible role for vitamin D deficiency in sleep disturbances (Bellia et al. 2013). These results are corroborated by intervention studies, but remain controversial (Huang et al. 2013; Majid et al. 2018). These results were found in a previous study with mining workers in the same region as this study; when evaluating sleep using the gold standard method, polysomnography, the workers with hypovitaminosis D had lower sleep efficiency, an increased microarousal index, and lower arterial oxygen saturation after adjusting for seasonality, age, and body fat (Menezes-Júnior et al. 2021). These workers had routines similar to confinement during the COVID-19 pandemic, since they were off-highway truck drivers and spent most of their time in machines inaccessible to sunlight (Menezes Júnior et al. 2021).

An additional variable associated with poor sleep quality in our study was the weight change during the pandemic. Individuals who reduced up to 5.0% of their body weight during the pandemic had a 66% greater chance of having poor sleep quality (OR= 1.66; 95%CI 1.01-2.76), and those who gained 5.0% of their body weight had a 90% greater chance (OR= 1.90; 95%CI 1.08-3.34). Weight loss, when intentional, especially in obese individuals, can be of great use in improving sleep quality (Hargens et al. 2013). However, unintentional weight loss may be related to increased physical and emotional stress, or food supply and demand. A systematic review conducted between July 2020 and February 2021, with 41 studies and 469,362 total participants, found that during the pandemic, 11.1%-32.0% of individuals had experienced weight loss (Khan et al. 2021). For some people, the lockdown provided more time to cook and eat better, but the majority of people suffered from malnutrition and weight loss because of inflated food prices and food insecurity. In Brazil, more than half of the households (59.4%) were food insecure during the pandemic (Galindo et al. 2021). Not eating enough food of adequate quantity and quality has health impacts, such as poor mental health and increased likelihood of diseases (Galindo et al. 2021), increasing the chances of poor sleep quality and higher vulnerability to COVID-19. Furthermore, individuals in above-normal stressful situations, such as those caused by the risk of SARS-CoV-2 contamination and social isolation, who have poor sleep quality, are at greater risk of depressive symptoms (Barros et al. 2019).

In addition, pandemic confinement was associated with weight gain in 7.2–72.4% of participants in a systematic review of 41 studies (Khan et al. 2021). Excess weight interferes with sleep quality in several aspects, by anatomical factors such as airway obstruction, or by inflammatory factors such as increased cytokines, which can induce sleep disturbances by altering the sleep-wake rhythm (Muscogiuri et al. 2019). Furthermore, there is a strong association that poor sleep quality may increase the risk of obesity, as demonstrated in longitudinal studies, such as in a cohort of 83,377 Americans, in which among men and women who were not obese at baseline, participants who reported less than five hours of sleep per night had an approximately 40% higher risk of developing obesity than those who reported seven to eight hours of sleep (for men, OR= 1.45, 95% CI: 1.06, 1.99; for women, OR= 1.37, 95% CI: 1.04, 1.79) (Xiao et al. 2013).

Regarding health variables, we found that individuals with at least one symptom of COVID-19 (fever, sore throat, cough, dyspnea, tachycardia, diarrhea, nausea, anosmia, ageusia, fatigue, and pain) in the 15 days before the study were twice as like likely to have poor sleep quality (OR=1.94; 95%CI 1.25-3.01). Insufficient sleep impairs the immune system and decreases resistance to viral diseases, increasing the risk of developing symptoms such as cough, nasal congestion, and catarrh, which cause discomfort and can disrupt sleep (Asif, Iqbal, and Nazir 2017). Common colds have symptoms similar to those of COVID-19. Cohen et al. conducted a clinical study assessing sleep for 14 days and administered nasal drops containing rhinovirus. They found that participants with less than seven hours of sleep were 2.94 times more likely to develop a cold than those with eight hours or more of sleep (95%CI: 1.18-7.30). In addition, sleep efficiency was associated with the total flu syndrome symptom score (β = -1.51 [SE = 0.40], p < 0.001), and people with flu symptoms have more difficulty initiating and sustaining sleep (Cohen et al. 2009).

Unfortunately, the fear and uncertainty caused by the pandemic, the threats to survival, among other things, are one of the main problems encountered during the pandemic and have greatly influenced the quality of life and mental health (Brooks et al. 2020). Of all the factors evaluated in our study, we found that anxiety and living alone were the most strongly associated with poor sleep. Individuals with anxiety or living alone were approximately twice as likely to have poor sleep quality.

Pandemic contexts and social isolation affect the population in many dimensions of living conditions and health status, particularly concerning the mental health component. In Brazil, 52.6% (95% CI 51.2; 54.1) of Brazilians frequently reported feeling anxious or nervous (Barros et al. 2020). Anxiety, especially generalized anxiety disorder, has been described as one of the most important consequences of sleep deprivation (Cox and Olatunji 2016). Thus, studies suggest that sleep disturbance aggravates anxiety disorder, but the causal role of sleep disturbance is still unclear (Cox and Olatunji 2016). A study during the first weeks of the lockdown in Italy found that lower sleep quality was directly related to days spent at home in confinement, with mental health playing an important role in mediating sleep quality (Casagrande et al. 2021). A systematic review and meta-analysis of 345,270 participants from 39 countries found consistent results regarding the association between sleep quality and psychological distress. The corrected pooled estimated prevalence of sleep problems was 18% among the general population and was positively associated with anxiety (Fisher’s z-score of 0.48; 95% CI: 0.41 to 0.54) (Alimoradi et al. 2021).

Psychological impact during a pandemic period is common and expected, as demonstrated by Brooks et al. (2020), who evaluated previous epidemics. The main psychological stressors were duration of quarantine, fear of infection, feelings of frustration and annoyance, inadequate information about care for the disease, financial losses, and stigma associated with the disease (Brooks et al. 2020). During the current COVID-19 pandemic, anxiety and sleep problems appear to have been common and affected a wide range of individuals and groups (Leão and Perelman 2018).

In addition to the previously mentioned factors, we also found that participants who had the co-occurrence of two factors associated with poor sleep quality had an increased chance of having impaired sleep quality. These results are important because the social and health context caused by the pandemic makes many subjects vulnerable to the co-occurrence of factors that negatively interfere with sleep quality. In this context, vitamin D insufficiency and weight gain, for example, are very related factors that can occur simultaneously (Bellia et al. 2013; Holick 2007). Therefore, the co-occurrence of these factors can increase the chances of poor sleep quality, as we have shown. To the best of our knowledge, this is the first study to evaluate the co-occurence of factors associated with poor sleep quality during the COVID-19 pandemic.

Insufficient sleep directly impacts the immune system and exponentially increases the chances of illness. Thus, we found a high prevalence of poor sleep quality during the COVID-19 pandemic, with several associated factors. Sleep quality may have been influenced by the COVID-19 pandemic and the government actions taken to contain it. Brazil is one of the countries with the highest number of deaths and lowest percentage of the population vaccinated, with vaccination expected to be completed only in December 2021 (MODCOVID19 2021), almost two years after the first coronavirus case in the country.

Sleep is an important factor to consider in a pandemic, given its interfaces with numerous other health conditions, as well as an improved immune response in the face of an opportunistic infection (Irwin 2015). Thus, a health emergency such as the one we are experiencing should be accompanied by adequate social support programs to mitigate the psychological, social, and economic effects, promoting a better situation to face a troubled period such as this.

The main limitations of this study are the variables obtained by self-report, which can lead to underestimation of risk behaviors or overestimation of protective behaviors, due to differences in the perception of each individual about the pandemic and associated factors. However, the assessment of sleep quality needs to be performed subjectively, since it considers intrinsic factors to the individuals’ perception of their sleep. Furthermore, the sample design brings robustness to the study and favors the analysis of the COVID-19 scenario in the two municipalities of the Iron Quadrangle region. Thus, this study allows us to evaluate the relationship between the quality of sleep and factors related to the pandemic, providing subsidies for decision making, in a chaotic socio-sanitary and epidemiological context, to reduce the worsening of health conditions.

## Data Availability

The data that support the findings of this study are available on request from the corresponding author [LAAM]. The data are not publicly available due to containing information that could compromise the privacy of research participants.

## Contributorship

GLLM, LGL, ACSA, JCCC, and ALM contributed to the conception and design of the work, to the acquisition, analysis, and interpretation of data, and to the draft of the manuscript. LAAMJ, LGL, ACSA, and ALM contributed to the analysis of the results and the writing of the manuscript. All authors have approved the submitted version.

## Acknowledge

The authors acknowledge the Federal University of Ouro Preto (UFOP) and the Research and Education Group in Nutrition and Collective Health (GPENSC) for their support and incentive. And also the support of the Municipal Health Secretariats of the municipalities evaluated in the study.

